# Changes in the Health of Adolescent Athletes: A Comparison of Health Measures Collected Before and During the CoVID-19 Pandemic

**DOI:** 10.1101/2021.01.12.20248726

**Authors:** Timothy A. McGuine, Kevin Biese, Scott J. Hetzel, Labina Petrovska, Stephanie Kliethermes, Claudia L. Reardon, David R. Bell, M. Alison Brooks, Andrew M. Watson

## Abstract

**Context:** In the spring of 2020, schools closed to in-person teaching and sports were cancelled to control the transmission of CoVID-19. The changes that took place to the physical and mental health among young athletes during this time remain unknown, however.

**Objective:** Identify changes in the health (mental health, physical activity and quality of life) of athletes that occurred during the CoVID-19 pandemic.

**Design:** Cross sectional study.

**Setting:** Sample recruited via social media.

**Patients or Other Participants:** 3243 Wisconsin adolescent athletes (age=16.2±1.2 yrs., female=58% female) completed an online survey in May 2020 (DuringCoVID-19). Health measures for this cohort were compared with previously reported data for Wisconsin adolescent athletes (n=5231, age=15.7±1.2, 47% female) collected in 2016–2018 (PreCoVID-19).

**Main Outcome Measure(s):** Demographic information included: sex, grade and sports played. Health assessments included the Patient Health Questionnaire-9 Item (PHQ-9) to identify depression symptoms, the Pediatric Functional Activity Brief Scale (PFABS) for physical activity, and the Pediatric Quality of Life Inventory 4.0 (PedsQL) for health related quality of life (HRQoL). Univariable comparisons of these variables between groups were made via t-tests or chi-square tests. Means and 95% confidence intervals (CI) for each group were estimated by survey weighted ANOVA models.

**RESULTS:** Compared to PreCoVID-19, a larger proportion of the During-CoVID-19 participants reported rates of moderate to severe levels of depression (9.7% vs 32.9%, p<0.001). During-CoVID-19 participants reported 50% lower (worse) PFABS scores (mean:12.2 [95%CI: 11.9, 12.5] vs 24.7 [24.5, 24.9] p<0.001) and lower (worse) PedsQL total scores compared to the PreCoVID-19 participants (78.4 [78.0, 78.8] vs. 90.9 [90.5, 91.3] p<0.001).

**CONCLUSIONS:** During the CoVID-19 pandemic, adolescent athletes reported increased symptoms of depression, decreased physical activity and decreased quality of life compared to adolescent athletes in previous years.

**Key points:**

1. Adolescent athletes during CoVID-19 were three times more likely to report moderate to severe symptoms of depression compared to data collected prior to CoVID-19.
2. Adolescent athletes during CoVID-19 reported significantly lower physical activity and quality of life scores compared to high school athletes prior to the CoVID-19 pandemic
3. Post CoVID-19 policies should be implemented to improve the health of adolescent athletes in the US.

## INTRODUCTION

An estimated 8.4 million US high school students participate in interscholastic athletics.^1^ Adolescent sport participation is recognized to have profound positive influences on the health and well-being of adolescent students as evidenced by higher academic achievement, greater levels of healthy physical activity and decreased levels of anxiety and depression compared to students who do not participate in athletics.^2-5^ Additionally, high school sport participation is one of the most important factors for life-long physical activity and health.^6-12^

During the winter and spring of 2020, CoVID-19, the disease caused by the novel SARS-CoV-2 coronavirus, reached pandemic proportions throughout the world. Across the US, schools were closed and athletic seasons were cancelled to slow the spread of the disease.

Experts have suggested that efforts to limit the spread of CoVID-19 may have profound societal, economic, and psychosocial consequences for students and need to be further studied.^13-17^

A recent study reported that female athletes, athletes in grade 12, team sport participants and athletes from areas with higher % of the population under age 18 in poverty reported lower levels of physical activity, greater symptoms of anxiety and depression, and lower HRQoL in May of 2020 during the nationwide shutdown.^18^ Describing the prevalence of mental health disorders during the CoVID-19-related restrictions is useful, but does not fully describe how specific populations of athletes have been impacted. We are not aware of research to date that has specifically documented mental and physical health *changes* in adolescent athletes during the pandemic. Quantifying these changes will allow health care providers to implement strategies to improve the health of adolescent athletes moving forward.^25^

An estimated 93,000 (33%) of the 284,000 Wisconsin high school students compete in interscholastic sports each year.^1^ At the start of the 2019/2020 school year, 478 Wisconsin schools offered interscholastic athletics and fielded teams competing in fifteen to twenty sports.^19^

These student athletes typically competed on one to three athletic teams throughout the school year (fall, winter and spring sport seasons), but many also continued to train with their teammates when their sport is not in season.^20,21^ Over half of these student athletes also competed on club sport teams not affiliated with their school during the school year.^22,23^

In March 2020, Wisconsin government and health officials mandated a two-week state-wide school closure to slow the spread of CoVID-19. The closure order was eventually extended twice to run through the end of the school year.^24^ While high school students’ academic work moved online, all in-person teaching and interscholastic athletics for students in grades 9–12 were cancelled. Additionally, adolescent club sport teams were restricted from participating during this time. While necessary to slow the community spread of the virus, the CoVID-19 mitigation strategies may nonetheless have negative effects on the physical and mental health of adolescents.^26^

Quantifying the changes to the health of adolescent athletes that occurred during the CoVID-19 pandemic is critical to understand how these issues impacted athletes in order to to improve management in the future.^27-29^ This information will allow sports medicine providers, school administrators and health care policy experts to implement strategies to improve the short-term and long-term mental and physical health of adolescent athletes in the months and years to come as we transition from the CoVID-19 pandemic.

The purpose of this study was to measure changes in the mental and physical health of Wisconsin adolescent athletes manifested during the CoVID-19 related school closures and sport cancellations. To measure these changes, we compared athlete self-report data on depression, physical activity and HRQoL that we collected during-CoVID-19 (DuringCoVID-19) with Wisconsin data from a series of previous investigations carried out by the study team during normal high school sport operations in the years 2015-2018 (PreCoVID-19).^30-34^ We hypothesize that adolescent student athletes will report significantly worse mental health, as well as lower quality of life and physical activity scores during the CoVID-19 pandemic shutdown, than has previously been reported prior to CoVID-19.

## METHODS

This study was approved by the University of Wisconsin-Madison Health Sciences Institutional Review Board in April of 2020. Wisconsin adolescent athletes (male and female, grade: 9–12, age:13-19) were recruited via social media (Facebook, Twitter) to participate in the study by completing an anonymous online survey in May 2020. To ensure widespread distribution and recruitment for the study, the study team provided the social media links to sports medicine provider colleagues across Wisconsin, the Wisconsin Interscholastic Athletic Association (WIAA) and the Wisconsin Athletic trainers’ Association (WATA), which provided the links to high school athletes throughout Wisconsin.

The survey was composed of 69 items and included a section to describe the demographics for the participant, which was followed by 3 segments that included validated instruments used to measure depression symptoms, physical activity and HRQoL in adolescents. Demographic information included items for the participants’ age, grade, school size, school funding (private or public) and zip code as well as a list of all the high school and club (not affiliated with their school) sports in which they competed during the previous 12 months. The remainder of the survey consisted of an assessment of mental health, physical activity level and HRQoL.

### Mental health

The Patient Health Questionnaire-9 Item (PHQ-9) survey was used to evaluate depression symptoms. This 9-item questionnaire asks participants to rate the frequency of depression symptoms experienced in the past two weeks.^35^ The PHQ-9 scores range from 0-27 with a higher score indicating a greater level of depression symptoms. The PHQ-9 has demonstrated high sensitivity and specificity for depression screening in adolescent patients aged 13 to 17 years. In addition to the total score, PHQ-9 categorical scores of 0–4, 5–9, 10–14, 15–19 and ≥ 20 correspond to minimal or none, mild, moderate, moderately severe and severe depression symptoms, respectively.^36,37^

### Physical Activity

Physical activity level was assessed with the Hospital for Special Surgery Pediatric Functional Activity Brief Scale (PFABS).^38,39^ This validated 8-item instrument was designed to measure the physical activity of children between 10 and 18 years old for the past month. Scores range from 0 to 30 with a higher score indicating greater physical activity.^38,39^

### Health Related Quality of Life

HRQoL was measured with Pediatric Quality of Life Inventory 4.0 (PedsQL). The 23-item PedsQL questionnaire assesses HRQoL for the previous 7 days. The PedsQL has been validated for use in children ages 2 to 18.^40,41^ Physical and psychosocial subscale summaries, as well as the total scores, range from 0 to 100 with a higher score indicating greater HRQoL. The PedsQL survey has been used to measure HRQoL in samples of healthy and injured adolescent athletes.^42^

### Statistical analyses

Statistical analyses were performed for participants who provided a valid, complete survey. Participants were excluded if they did not complete the entire survey, were not in grades 9 -12, or indicated they did not participate in club or interscholastic sports within the previous 12 months.

Demographic variables and individual sport participation were summarized (mean (SD) or N (%)) overall and by study group (PreCoVID-19 athlete cohort and During-CoVID-19 athlete cohort) stratified by sex. Univariable comparisons of these variables between groups were made via t-tests or chi-square tests. Means and 95% confidence intervals (CI) for each group were estimated by survey weighted ANOVA models separately for depression, physical activity and quality of life measures. A weighted ordinal logistic regression (OLR) model was used to estimate the percentages of level of depression (PHQ-9) for each group. A group by sex interaction was modeled for all sex-specific estimates.

Sample survey weights were derived based on sport and sex using the 2018-2019 Wisconsin high school sport participation statistics compiled annually by the National Federation of High Schools (NFHS).^1^ Survey weights for multiple sport athletes were based on the most common sport played. Inverse propensity score weights (IPSW) were also calculated to account for design-based differences in the two groups.^43^ Propensity score weights were assigned using logistic regression which was weighted based on the sample survey weights. Group assignment was the dependent variable, and age, sex, and high school individual sport status were predictor variables.

The PHQ9, PFABS and PedsQL scores were compared with historical data of Wisconsin adolescent athletes collected during normal school operations in the years 2015–2018.^25-29^Final models comparing the During-CoVID-19 group to the PreCoVID-19 group included weights as the multiplication of the sampling weights and IPSWs.^44^

## RESULTS

A total of 3,243 Wisconsin adolescent athletes (age = 16.2±1.2 yrs., female = 58%) completed the survey in May 2020 (During-CoVID-19) with responses obtained from participants residing in 71 (99%) of the 72 counties in Wisconsin. Due to the convenience sampling design, information regarding the response rate was unavailable. A total of 2,730 (84%) participants indicated they attended a publicly funded school with the median [IQR] school enrollment = 802 [401, 1248]. Nine hundred seventy (29.9%) competed in a single sport while the remaining 2,273 (70.1%) competed in two or more sports for their school in the prior 12 months. During-CoVID-19 participants were most likely to compete in track (32.8%), basketball (29.1%) and volleyball (21.2%). In addition to competing for their high school teams, 1,962 (60.5%) competed as an individual or as part of a club team outside their school setting.

The PreCoVID-19 cohort consisted of 5,231athletes (age = 15.7±1.2, female = 65%) that were enrolled in cohort and longitudinal studies in 2015-2018.^30-34^ A summary of the number of athletes by sex, age and grade for both the During-CoVID-19 and PreCoVID-19 cohorts are found in Table 1. The PreCoVID-19 participants resided in 56 (78%) of Wisconsin counties. A total of 4,399 (84%) of these participants attended a publicly funded school with the median [IQR] school enrollment = 654 [299, 1017]. The PreCoVID-19 participants were most likely to compete in volleyball (55.7%), basketball (43.2%) and football (23.3%). A summary of the number of athletes and percentages of the sports played for both the During-CoVID-19 and PreCoVID-19 cohorts are found in Supplementary Table 1.

**Table 1.** Comparison of Depression, Physical Activity and HRQoL Scores for Adolescent Athletes Prior to the CoVID-19 Pandemic (PreCoVID-19) and During the CoVID-19 Pandemic (During-CoVID-19).

| Variable | PreCoVID-19<br>All<br>Participants<br>(n = 5231) | During-CoVID-19<br>All<br>Participants<br>N = 3243 | P value | PreCoVID-19<br>Females<br>(n = 3402) | During-CoVID-19<br>Females<br>(n = 1877) | P value | PreCoVID-19<br>Males<br>(n = 1829) | During-CoVID-19<br>Males<br>(n = 1366) | P value |
| --- | --- | --- | --- | --- | --- | --- | --- | --- | --- |
| Sex – Male<sup>^</sup> | 1829 (35.0%) | 1366 (42.1%) | < 0.001 | -- | -- |  | -- | -- |  |
| Age<sup>^</sup> | 15.7 (1.1) | 16.2 (1.2) | < 0.001 | 15.6 (1.1) | 16.1 (1.2) | < 0.001 | 15.8 (1.2) | 16.2 (1.2) | < 0.001 |
| Grade<sup>^</sup> |  |  | < 0.001 |  |  | < 0.001 |  |  | 0.003 |
| 9 | 1763 (33.7%) | 862 (26.6%) |  | 1177 (34.6%) | 499 (26.6%) |  | 586 (32.0%) | 363 (26.6%) |  |
| 10 | 1388 (26.5%) | 845 (26.1%) |  | 937 (27.5%) | 509 (27.1%) |  | 451 (24.7%) | 336 (24.6%) |  |
| 11 | 1153 (22.0%) | 848 (26.1%) |  | 720 (21.2%) | 470 (25.0%) |  | 433 (23.7%) | 378 (27.7%) |  |
| 12 | 927 (17.7%) | 688 (21.2%) |  | 568 (16.7%) | 399 (21.3%) |  | 359 (19.6%) | 289 (21.2%) |  |
| PHQ9 Total Score ** | 3.3 (3.1, 3.5) | 8.0 (7.8, 8.2) | < 0.001 | 3.6 (3.4, 3.8) | 8.2 (7.9, 8.5) | < 0.001 | 2.6 (2.2, 2.9) | 7.8 (7.5, 8.1) | < 0.001 |
| PFABS Total Score * | 24.7 (24.5, 24.9) | 12.2 (11.9, 12.5) | < 0.001 | 24.5 (24.2, 24.7) | 11.4 (11.1, 11.8) | < 0.001 | 25.7 (25.3, 26.1) | 13.9 (13.4, 14.4) | < 0.001 |
| PedsQL *** |  |  |  |  |  |  |  |  |  |
| Physical Summary | 91.7 (91.3, 92.1) | 82.6 (82.2, 83.0) | < 0.001 | 89.6 (89.1, 90.0) | 80.9 (80.5, 81.4) | < 0.001 | 96.6 (95.9, 97.3) | 86.1 (85.4, 86.8) | < 0.001 |
| Psychosocial Summary | 90.4 (89.9, 90.8) | 76.2 (75.8, 76.6) | < 0.001 | 90.1 (89.6, 90.6) | 75.0 (74.5, 75.5) | < 0.001 | 90.9 (90.2, 91.7) | 78.6 (77.9, 79.3) | < 0.001 |
| PedsQL Total | 90.9 (90.5, 91.3) | 78.4 (78.0, 78.8) | < 0.001 | 90.1 (89.6, 90.6) | 75.0 (74.5, 75.5) | < 0.001 | 92.9 (92.2, 93.6) | 81.2 (80.6, 81.9) | < 0.001 |

\* Historical group N=1002; \*\* Historical group N = 2156; \*\*\* Historical group N = 5231
<sup>^</sup> Reported as n (%) for Sex and Grade, mean (SD) for Age,
Total scores for the PFABS, PHQ-9 and PedsQL reported as the sample weight \* inverse propensity score weighted mean (95% CI)

### Mental health

The number of females that reported a mild level of depression in the During-CoVID-19 athletes was 43% higher compared to the PreCoVID-19 females (During-CoVID-19 = 35%, PreCoVID-19 = 24%, p <0.001). Females in the During-CoVID-19 group reported a three-fold increase in moderate, moderately severe or severe levels of depression compared to the PreCoVID-19 athletes (During-CoVID-19 = 37%, PreCoVID-19 = 11%, p <0.001). The prevalence of depression levels for females in both the PreCoVID-19 and During-CoVID-19 groups are found in Figure 1. The number of males in the During-CoVID-19 group that reported a mild level of depression was 130% higher than compared to PreCoVID-19 males (p (During-CoVID-19 = 35%, PreCoVID-19 = 15%, p <0.001). The proportion of males in the During-CoVID-19 group that reported moderate, moderately severe or severe levels of depression was 4.5 times higher than males in the PreCoVID-19 group (During-CoVID-19 = 27%, PreCoVID-19 = 6%, p <0.001). p <0.001). The prevalence of depression levels for males in the During-CoVID-19 and PreCoVID-19 group is found in Figure 2.

**Figure 1.**
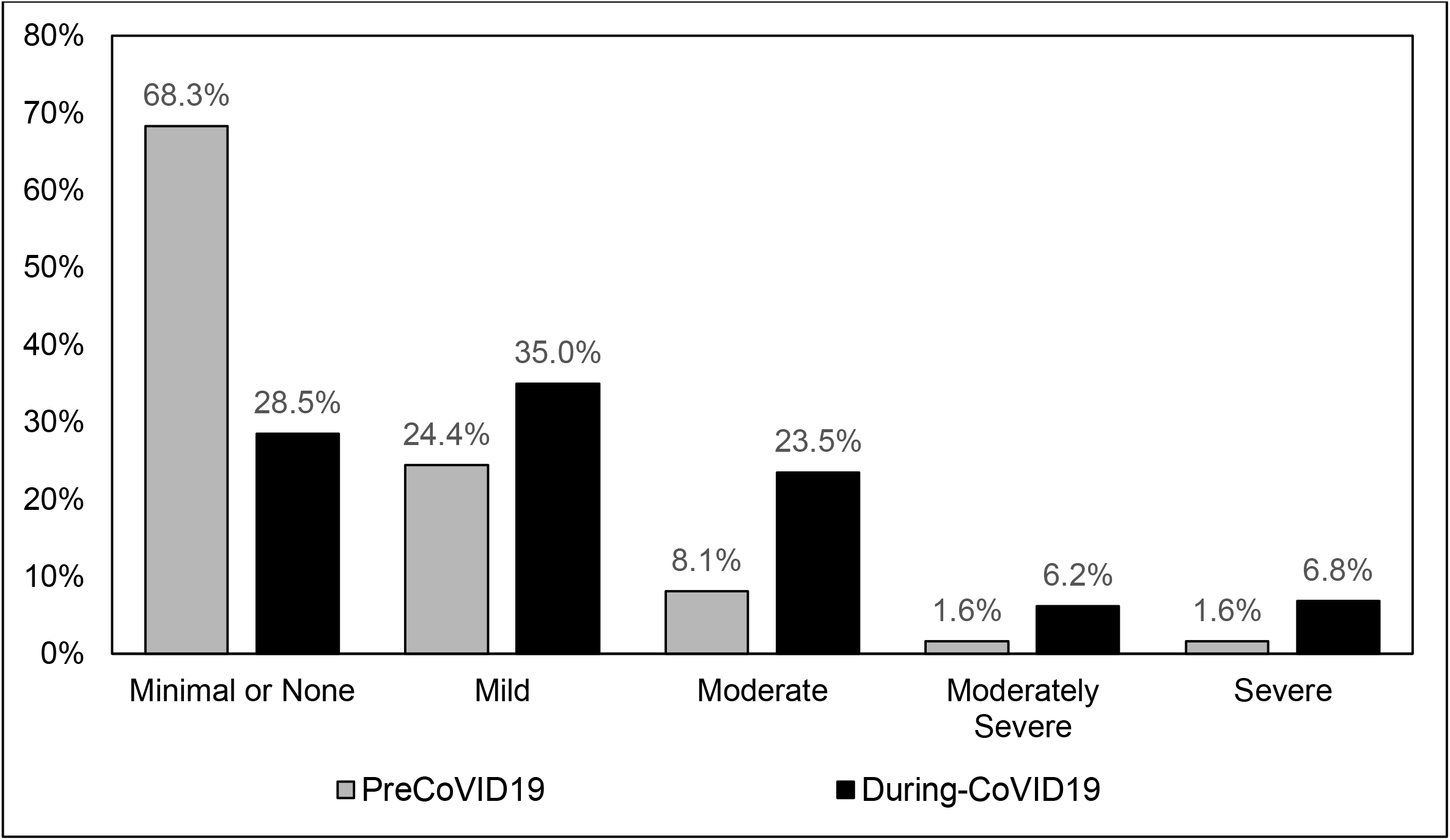
Prevalence of Depression Symptoms for Female Adolescent Athletes Prior to the COVID-19 Pandemic (PreCoVID-19) and in During the CoVID-19 Pandemic (DuringCoVID-19). PHQ9 categories are sample weight * inverse propensity score weighted % (95% CI)

**Figure 2.**
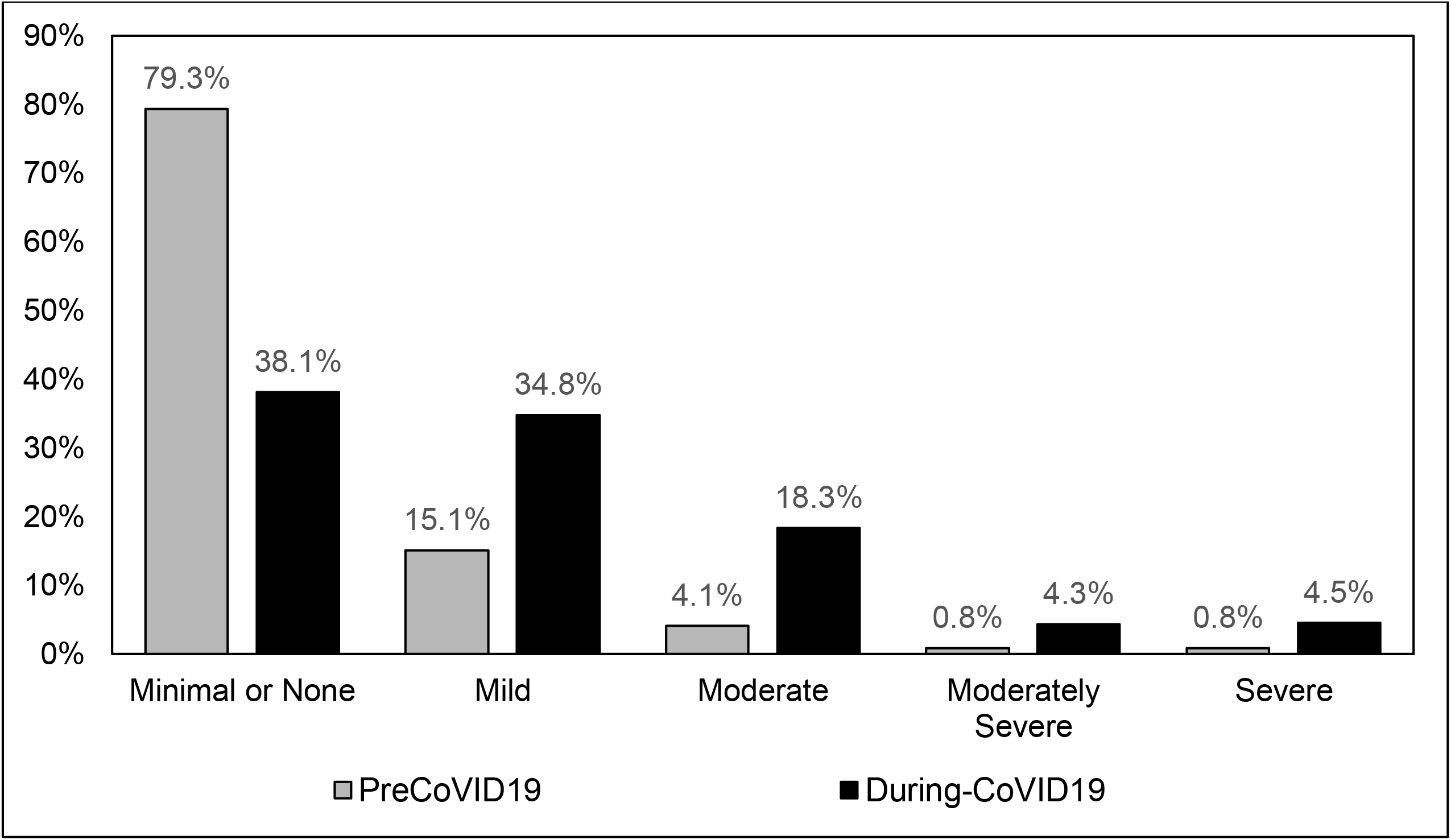
Prevalence of Depression Symptoms for Male Adolescent Athletes Prior to the COVID-19 Pandemic (PreCoVID-19) and in During the CoVID-19 Pandemic (DuringCoVID-19). PHQ9 categories are sample weight * inverse propensity score weighted % (95% CI)

Overall, the PHQ-9 scores for athletes During-CoVID-19 were 2.5 times higher (worse) than the athletes in the PreCoVID-19 group. The During-CoVID-19 females reported a (mean [95%CI]) PHQ-9 score that was over twice the PHQ-9 score for females in the PreCoVID-19 group (8.2 [7.9, 8.5] vs. 3.6 [3.4, 3.8]). Similarly, the During-CoVID-19 males had PHQ-9 scores that were three-times higher than the males in the PreCoVID-19 group (7.8 [7.5, 8.1] vs. 2.6 [2.2, 2.9]. The PHQ-9 scores for both groups are found in Table 1.

### Physical Activity

The total PFABS scores (mean [95%CI]) were lower (less physical activity) for athletes in the During-CoVID-19 group compared to the PreCoVID-19 group (12.2 [11.9.12.5] vs. 27.7 [24.5.24.9], p < 0.001). The During-CoVID-19 females reported PFABS scores that were more than 50% lower (worse) than the PreCoVID-19 females (11.4 [11.1, 11.8] vs. 24.5 [24.2, 24.7], p < 0.001). Similarly, males in the During-CoVID-19 group reported PFABS scores approximately 50% lower compared to athletes in the PreCoVID-19 group (13.9 [13.4, 14.4] vs. 25.7 [25.3, 26.1], p < 0.001). The PFABS scores for both groups are found in Table 1.

### Health Related Quality of Life

Higher (better) PedsQL physical summary, psychosocial summary, and PedsQL total scores were reported for athletes in the PreCoVID-19 group compared to the athletes in the During-CoVID-19 group (p <0.001). The PreCoVID-19 physical health summary scores (mean [95%CI]) for females were 11% higher than those reported in the During-CoVID-19 group (89.6 [89.1, 90.0] vs. 80.9 [80.5, 81.4], p < 0.001). Similarly, the PreCoVID-19 physical health summary scores for females were 20% higher than the scores reported in the During-CoVID-19 group (90.1 [89.6, 90.6] vs. 75.0 [74.5, 75.5], p < 0.001), while the PreCoVID-19 total PedsQL scores for females were 17% higher than the scores reported in the During-CoVID-19 group (90.0 [89.5, 90.4] vs. 77.1 [76.6, 77.6], p < 0.001).

The PreCoVID-19 physical health summary scores (mean [95%CI]) for males were 12% higher than the scores reported by athletes in the During-CoVID-19 group (96.6 [95.9, 97.3] vs. 86.1 [85.4, 86.8], p < 0.001). Similarly, the PreCoVID-19 physical health summary scores for males were 14% higher than the scores reported by males in the During-CoVID-19 group (90.9 [90.2, 91.7] vs. 78.6 [77.9, 79.3], p < 0.001). The PedsQL total scores reported by the males in the PreCoVID-19 group were also 14% higher than scores for males in the During-CoVID-19 group (92.9 [92.2, 93.6] vs. 81.2.1 [80.6, 81.9], p < 0.001). The PedsQL scores for both groups are found in Table 1.

## DISCUSSION

The primary findings of this study are that the mental health, physical activity and HRQoL scores for adolescent athletes were significantly worse during the CoVID-19 pandemic than scores reported by adolescent athletes in prior years. To our knowledge there are no existing data that demonstrates mental health, physical activity and HRQoL *changes* for adolescent athletes immediately following the cancellation of school and athletic seasons during the CoVID-19 pandemic. It has been suggested that the negative psychosocial effects of CoVID-19 related mitigation measures may be manifested in additional health care utilization and spending in the coming months and years.^25,26^ The results of this study can be used to inform stakeholders of the results of quarantine restrictions and how they negatively impact the health of adolescent athletes.

### Mental Health

The levels of depression symptoms and total symptom scores reported (as measured by the PHQ-9) in the During-CoVID-19 cohort were significantly higher than our PreCoVID-19 athletes. Additionally, our During-CoVID-19 cohort had higher depression symptoms than previous studies of adolescents that observed severe depression symptoms in approximately 2% of adolescents.^45^ While the prevalence of moderate to severe depressive symptoms was greater among females than males in the During-CoVID-19, both males and females demonstrated dramatically higher rates compared to data collected before the CoVID-19 pandemic.^45^ The depression data reported for participants in the During-CoVID-19 group demonstrate a significant mental health burden among this population of young athletes.

The decrease in mental health is multifaceted and complex and may be related to the decreased socialization, increased family strain, and reduced access to support services.^46^ In Wisconsin, many families self-isolated or severely limited social contact for several weeks to several months in the Spring of 2020. Previous studies have demonstrated that quarantines can negatively impact mental health.^46,47^ Schools play an important role in providing access to mental health services for disadvantaged populations, and health care providers, parents, and policy-makers should be mindful of the mental health strain the current pandemic is placing on adolescents.^48^

### Physical Activity

Our study is the first in the United States to demonstrate a significant decrease in physical activity levels in adolescents during the CoVID-19 pandemic. While male athletes reported slightly higher levels of physical activity than females, the decrease in both males and females during the CoVID-19 restrictions was similar, with current levels of physical activity approximately 50% lower than levels reported in a similar population prior to CoVID-19 reported by Fabricant.^38^ Physical activity is known to have a beneficial effect on a wide range of health outcomes in adolescents, including sleep, academic success, well-being, and mental health.^49-51^ Therefore, it is possible that the identified decrease in mental health in our participants may be at least partly due to the removal of the positive effects that physical activity has for adolescents.

Childhood obesity was a health care crisis prior to the COVID-19 pandemic that is increasing in the US and is projected to become worse due to the pandemic.^28,29,52^ A decrease in physical activity and increase in stress may increase the risk of severe COVID-19 cases in obese children.^29^ Decreased physical activity in adolescents may also have longer-term negative effects and implications in terms of increased risk for obesity and cardiometabolic disease if these levels remain low for prolonged periods.^53^ Chronically low levels of physical activity may also compound the mental health consequences of the current crisis.^49,50^ Further, there is evidence that sport participation during high school is related to improved health and well-being throughout adulthood.^7-11^ As time spent being physical active during the school day has decreased, organized sports have been proposed as a means to stabilize long-term physical activity in adolescents and improve adult physical activity.^49^ Though returning adolescents to organized sports is a complex issue and requires careful consideration, stakeholders should consider the promotion of physical activity a top priority during the CoVID-19 pandemic.

### Health Related Quality of Life

Several studies have shown that individuals with increased physical activity and or interscholastic sport participation report higher HRQoL scores compared to inactive adolescents and high school non-athletes.^42,54^ Therefore, it is not surprising that HRQoL scores decreased in the During-CoVID-19 cohort compared to the PreCoVID-19 athletes. Interestingly, the overall PedsQL scores observed for the During-CoVID-19 athletes are similar to scores recorded in the general adolescent population (during the pandemic?) but higher than scores reported in adolescents with chronic disease conditions (during the pandemic?).^40,55^

In our study, females reported higher levels of depression symptoms as well as lower physical activity and HRQoL scores than males. This is consistent with previous research on depression, physical activity and HRQoL in adolescents.^36-40,50,52,56^ Therefore it is unlikely that the sex differences in depression symptoms and HRQoL scores we noted are attributable to the CoVID-19 pandemic directly. Nonetheless, we found dramatic reductions in HRQoL for both males and females during the COVID-19 shutdown, and researchers should continue to monitor HRQoL in adolescents as separation from peers and sports continue during restrictions initiated to limit the spread of CoVID-19.

### Limitations

This study has several limitations. First, the data provided were self-reported from online surveys and not the result of a clinical examination conducted by a health care provider. Nonetheless, our findings include a large sample of athletes and align with reports from experts who have stated that CoVID-19 will impact the health of youth populations.^13,16,17,25,26^ Second, we acknowledge that there may be a response bias for participants. We cannot know for certain if the sample is representative of all Wisconsin adolescent athletes or biased towards athletes who were more likely to respond if they experienced the most profound impacts on their health. Third, due to the survey delivery method, our sample may be biased toward athletes from higher socioeconomic families with easy access to internet services and social media platforms. We could not eliminate this bias, but we did collect results from 99% of Wisconsin counties, which span a wide range of socioeconomic levels. Further, we utilized medical colleagues as well as the Wisconsin Interscholastic Athletic Association and the Wisconsin Athletic Trainers’ Association to publicize the study and urge all adolescent athletes to participate. In addition, we used recent high school sport participation statistics from Wisconsin schools to calculate sample weights to mitigate the potential effects of selection bias. This increased the representativeness of our results to the entire adolescent athlete population in Wisconsin. Additionally, it is possible that other variables, such as participation in other co-curricular activities and missing school related events, were not accounted for in our study and could confound the results. Finally, our study did not include a control group consisting of non-athletes. Our existing data sets only include information on athletes, therefore we could only address potential changes over time (Pre-COVID to During-COVID) in athletic populations. We acknowledge it is very likely that the impacts observed in this sample of athletes would be observed across a wide spectrum of youth due the cancellations of most extracurricular activities.

## CONCLUSION

The physical activity and mental health of adolescent athletes worsened significantly during the CoVID-19 pandemic compared to PreCoVID-19 values. Our findings suggest that adolescent athletes have experienced dramatic increases in symptoms of depression along with subsequent decreases in physical activity and HRQoL during this time. Sports medicine and health policy experts should consider the potential negative mental and physical health effects that have become prevalent among adolescent athletes during the CoVID-19 pandemic, and stakeholders should consider systematic steps than can be implemented to reduce these negative changes in the months and years to come.

## Data Availability

The data can be made available by contacting the study PI

---

**Supplementary Table 1.** Distribution of Adolescent Sport Participants for the PreCoVID-19 and During-CoVID-19 Groups.

| **Sport** | **PreCoVID-19**<br>**All participants = 5,231**<br>**n (%)** | **PreCoVID-19**<br>**Females = 3,402**<br>**n (%)** | **During-CoVID-19**<br>**Females = 1,877**<br>**n (%)** | **PreCoVID-19**<br>**Males = 1,829**<br>**n (%)** | **During-CoVID-19**<br>**Males = 1,366**<br>**n (%)** |
| --- | --- | --- | --- | --- | --- |
| Baseball | 537 (10.3%) | 5 (0.1%) | 1 (0.1%) | 532 (29.1%) | 417 (30.5%) |
| Basketball | 2259 (43.2%) | 1590 (46.7%) | 502 (26.7%) | 669 (36.6%) | 437 (32.0%) |
| Cheer | 117 (2.2%) | 115 (3.4%) | 127 (6.8%) | 2 (0.1%) | 4 (0.3%) |
| Cross Country | 122 (2.3%) | 93 (2.7%) | 272 (14.5%) | 29 (1.6%) | 159 (11.6%) |
| Football | 1218 (23.3%) | 6 (0.2%) | 14 (0.7%) | 1212 (66.3%) | 621 (45.5%) |
| Golf | 30 (0.6%) | 24 (0.7%) | 73 (3.9%) | 6 (0.3%) | 90 (6.6%) |
| Gymnastics | 48 (0.9%) | 48 (1.4%) | 31 (1.7%) | 0 (0.0%) | 0 (0.0%) |
| Ice Hockey | 111 (2.1%) | 26 (0.8%) | 38 (2.0%) | 85 (4.6%) | 44 (3.2%) |
| Lacrosse | 91 (1.7%) | 16 (0.5%) | 19 (1.0%) | 75 (4.1%) | 15 (1.1%) |
| Soccer | 749 (14.3%) | 540 (15.9%) | 391 (20.8%) | 209 (11.4%) | 189 (13.8%) |
| Softball | 848 (16.2%) | 846 (24.9%) | 504 (26.9%) | 2 (0.1%) | 3 (0.2%) |
| Swimming | 28 (0.5%) | 22 (0.6%) | 86 (4.6%) | 6 (0.3%) | 40 (2.9%) |
| Tennis | 75 (1.4%) | 70 (2.1%) | 114 (6.1%) | 5 (0.3%) | 52 (3.8%) |
| Track | 1117 (21.4%) | 907 (26.7%) | 588 (31.3%) | 210 (11.5%) | 468 (34.3%) |
| Volleyball | 2916 (55.7%) | 2907 (85.4%) | 651 (34.7%) | 9 (0.5%) | 28 (2.0%) |
| Wrestling | 123 (2.4%) | 3 (0.1%) | 30 (1.6%) | 120 (6.6%) | 174 (12.7%) |
| Other<sup>\$</sup> | 62 (1.2%) | 55 (1.7%) | 94 (5.0%) | 7 (0.4%) | 51 (3.7%) |
| Total Sports Played<sup>^</sup> | 2.0 (0.8) | 2.1 (0.8) | 1.9 (0.8) | 1.7 (0.7) | 2.0 (0.8) |

<sup>^</sup>: reported as Mean (SD)
<sup>\$</sup> Other sports: Archery, Bowling, Field Hockey, Rugby, Snow Skiing (Alpine or Nordic), Power lifting, Trap Shooting, Ultimate
Frisbee

## Notes

### Competing Interest Statement

The authors have declared no competing interest.

### Funding Statement

No external funding was received for this study

### Author Declarations

This study (MR-HSIRB 2020-0554) was reviewed and approved by the Minimal Risk IRB (Health Sciences) at the University of Wisconsin

